# Development of A Migraine Trigger Measurement System Using Surprisal

**DOI:** 10.1101/2025.02.27.25322488

**Authors:** Dana P. Turner, Emily Caplis, Twinkle Patel, Timothy T. Houle

## Abstract

**Background:** Individuals who experience migraine continually seek to understand the causes, or “triggers,” of their attacks. Many triggers have been hypothesized, and it is not uncommon for individuals with migraine to be advised to consider a vast number of potential migraine triggers ranging from foods, weather influences, stress, mood states, certain behaviors, and sleep, among many others. Information-theoretic measures such as “surprisal” offer a novel approach to quantifying the unpredictability and diversity of trigger exposures on a single standardized scale.

**Objective:** This study aimed to quantify the within- and between-person variability of migraine trigger exposures using surprisal and entropy measures and to evaluate their potential for stratifying individuals based on trigger exposure patterns.

**Methods:** This longitudinal daily diary study included participants diagnosed with migraine who completed twice-daily electronic diaries reporting exposures to a range of potential headache triggers. Surprisal values were calculated to quantify the unexpectedness of individual trigger exposures, while entropy values captured overall variability in trigger domains such as sleep, mood, daily stressors, dietary behaviors, and environmental encounters.

**Results:** N = 109 individuals enrolled in the study and self-reported 187 different headache triggers for up to 28 days, resulting in 540,876 headache trigger measurements. Participants exhibited substantial heterogeneity in surprisal and entropy values across trigger domains, reflecting diverse patterns of exposure both within and between individuals. Morning measures of sleep and mood showed moderate entropy, while evening measures of dietary patterns and environmental encounters exhibited greater variability. A small number of principal components explained most of the variability in surprisal values, suggesting that only a few dimensions might offer the ability to characterize trigger exposure across the variables.

**Conclusions:** These findings reinforce the utility of surprisal measures for capturing nuanced patterns in the vast array of headache trigger data and support their potential as a measurement tool for stratifying trigger exposure either at the day or individual level.

## Introduction

Migraine headache disorders are widely prevalent and highly disabling.^1–3^ High costs and diminished quality of life are frequent effects of such disorders.^1,2^ Thus, individuals who experience migraine continually seek the causes, or “triggers,” of their attacks in an attempt to prevent attacks from occurring.^4^ Many such triggers have been hypothesized for millennia.^5^ Observational studies have provided much of what is known about headache triggers, but the majority of such studies have relied on simple checklists to assess whether individuals consider certain factors to be triggers. Further, despite recent advances in the understanding of pathophysiology,^6,7^ we still do not know what causes migraine attacks. It is not uncommon for individuals with migraine to be advised to consider a vast number of potential migraine triggers that range from foods, weather influences, stress, mood states, certain behaviors, sleep, and many others.

A potentially useful concept for studying the vast array of migraine triggers could be the amount of information contained in a trigger.^8^ Information theory has been used to quantify the amount of information in a variable^9^ by relating the amount of information to the uncertainty in observing the levels of a variable. The amount of information in a single trigger exposure can also be quantified and is called the “surprisal,” or the amount of surprise in observing the trigger.^10^ There is much more information contained in surprising events than in ones that are expected. For binary (present vs absent) triggers, surprisal is quantified as Surprisal = -log_2_(*p*), where the probability of experiencing the event is represented by *p*, and the resulting value is communicated in bits of information.^11^ Using information-theoretic measures, such as surprisal, has great potential for allowing the creation of a universal headache trigger measurement system in which the influence of a large and diverse set of headache trigger variables could be represented on the same scale.

The current study is based on our previous observations that different migraine triggers exhibit remarkably different amounts of daily information^8^ and that measuring the variation in the sum of “surprisals” exhibited by all trigger candidates has substantial value for measuring the total influence of experienced triggers.^10^ In this follow-up study, we aimed to examine the within-person and between-person variability in commonplace migraine trigger distributions using our previously delineated definition of surprisal. We hypothesized that, for the triggers under study, individualized (within-person) probability distributions exhibit variability and could be used to estimate total daily surprisal constituted by all observed trigger variables. We further hypothesized that surprisal distributions for headache triggers exhibit substantial between-person variability that could be used to stratify individuals based on variations in their trigger-exposure risk.

## Methods

This manuscript represents one of four pre-planned primary analyses of these data. This longitudinal daily diary study was conducted from April 2021 to December 2024 and was approved by the local Institutional Review Board. Participants were recruited from the local community using our institution’s online research recruitment platform, advertisements on public transportation, and flyers posted in the community. Interested potential participants were screened by telephone for eligibility criteria. Inclusion criteria were 1) Diagnosis of migraine headache disorder with or without aura that meets the International Classification of Headache Disorders, 3rd Edition, ICHD-3; 2) 4 to 14 headache days/month; and 3) Age of 18 to 65 years. Exclusion criteria were 1) Presence of a secondary headache disorder; 2) Chronic daily headache or medication over-use headache; 3) Recent change in nature of headache symptoms over last 6 weeks; 4) Unable to read or speak English at a 6^th^ grade level; 5) Any unmanaged Axis I psychotic disorder; 6) An active substance dependence issue (e.g., alcohol, marijuana) that interferes with data collection and headache activity; and 7) Pregnancy or planned pregnancy during the 28-day observation period.

Those found eligible were invited to an enrollment session, conducted either in person or virtually. Before beginning participation, each individual completed the informed consent process using the electronic informed consent functionality of the REDCap platform.^12^ Participants then began a series of enrollment questionnaires that included demographic information, headache characteristics, and the Migraine Disability Assessment (MIDAS)^13^ in REDCap. After completing the questionnaires, participants were instructed on the study procedures that they would be completing at home. These procedures included completing twice-daily diaries each morning and evening for 28 days. The diary entries were done in REDCap and took approximately 5 to 10 minutes to complete. Wearable data were also collected, but because the scale (i.e., moment to moment) differs greatly from the twice-daily assessments, it will be analyzed in a subsequent manuscript. Additional questionnaires were administered during enrollment and completion sessions but are not summarized in this manuscript.

### Daily Diary Items

The twice-daily electronic diaries were designed to capture the exposure to different putative migraine triggers. The diaries also contained items related to measures of headache activity and medication use. Due to the nature of the trigger constructs, a different set of triggers were assessed after waking (AM) and before bedtime (PM). The AM diaries assessed sleep behaviors,^14^ including duration, quality, nocturnal awakenings, bedtimes, and wake times. The AM diaries also assessed late-night meal patterns, the impact of weather, and mood state from the Profile of Mood States Short Form (POMS-SF^15^). The PM diaries also assessed mood state using the POMS-SF and included measures of daily stressors from the Daily Stress Inventory (DSI^16^). Major categories of commonly endorsed food and drink triggers^4^, environmental trigger encounters, caffeine intake, alcohol intake, balance symptoms, meal patterns, and weather impact were also assessed.

### Statistical Power Considerations

The hypotheses for this analysis are primarily descriptive. However, the originally intended sample size of N = 200 allowed for 95% confidence interval (CI) precision of +-4.16% for an observed proportion of 10%. Unfortunately, the COVID-19 pandemic affected the number of participants who could be enrolled. A research shutdown, a hiring freeze, and potential participants’ concerns about disease transmission made it difficult to enroll participants for an extended period of time. The final number of enrolled participants was N = 109. This number was considered sufficient to meaningfully evaluate the study’s hypotheses.

### Surprisal Methods

Understanding of the influence of potential headache triggers can be enhanced by concepts in information theory, which quantifies the “information” in a variable by measuring its uncertainty.^8,10^ Variables with higher variance convey more information on average. For instance, if a food item is consumed 25% of the time but caffeine is never consumed, the food item conveys some information due to its variability, while there is no information on caffeine consumption. This average information is captured by a measure called *information entropy*.^9^

Beyond average information, the information contained by a single event, or its “surprisal,” depends on its rarity. Surprising events carry more information than expected ones. For example, the surprisal of encountering a rare trigger (probability = 0.02, or occurring on only 2% of occasions) is high and can be quantified as −log_2_(p), expressed in bits (-log2[.02] = 5.6 bits). A fair coin toss results in 1 bit of surprisal (−log_2_[0.5] = 1 bit), while two consecutive heads yield 2 bits. This scale applies to events ranging from common (1 bit) to rare (e.g., rolling two dice and getting ones on both [“snake eyes”] twice in a row, ∼10 bits). Surprisal quantifies unexpectedness for binary variables and can extend to more complex probability distributions, including those for any headache trigger.

There are many ways that the surprisal of a trigger could be estimated for a single observed exposure. For example, the event could be surprising to the extent that it is rarely encountered by a population of individuals. Conversely, the surprisal of an event could be estimated based only on how surprising it is to *this* individual while ignoring the experiences of others. Additionally, the timing of the surprise can be weighted to reflect the assumption that recent exposures carry more significance than past ones (e.g., drinking coffee infrequently but recently is less surprising than drinking coffee infrequently with no recent occurrences). In the current analysis, surprisal was estimated at the level of the individual while ignoring the experiences of others using the following formula:

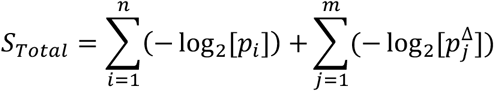

Where the sum of surprisal (S_Total_) is derived from the sum of the individual trigger exposure probabilities (p_i_) and added to the sum of the probability of observing a change in the individual trigger exposure 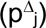, reflecting a difference between consecutive observations of the same measurement.

### Statistical Analysis

The analysis focused on descriptive and exploratory methods to characterize patterns of variability and covariance in headache trigger exposures summarized by entropy and surprisal. Descriptive statistics were calculated to summarize participant demographics, headache characteristics, and trigger-related variables. Continuous variables were summarized using medians and interquartile ranges (IQR), while categorical variables were presented as frequencies and percentages. Information entropy (i.e., average surprisal), individual surprisal values, and total surprisal (see Formula 1) were computed to quantify the variability and predictability of participants’ exposures to potential headache triggers. The entropy values for individual triggers and trigger domains were calculated to explore within- and between-person variability in trigger experiences, with results stratified by domains such as sleep, mood, daily stressors, dietary behaviors, and environmental encounters. To evaluate patterns of association between variables, we constructed separate AM and PM correlation matrices using Pearson correlation coefficients. Principal Component Analysis (PCA) was performed separately for AM and PM surprisal data to reduce dimensionality and identify latent patterns of variability across the trigger domains. To account for potential bias introduced by repeated measures nested within individuals, we used the **lme4** package to fit mixed-effects models and removed the participant-level variance from the surprisal values before conducting the PCA, The PCA used eigenvalue scree plots to determine the number of principal components needed to explain the majority of variance in the data. All statistical analyses were conducted using R 4.1, with results visualized using **ggplot2** and associated packages.

## Results

The study included 109 participants with a median age of 35 years [26.0, 46.0], of whom the majority were female (93.5%). Participants primarily identified as White (83.5%), followed by Asian (8.3%), Hispanic (10.2%), Black (5.5%), and American Indian or Alaska Native (2.8%). Most participants were single, 54.6%, while 40.7% were married. Regarding headache characteristics, all participants experienced migraine headache disorder as part of the inclusion criteria and reported a median headache frequency of 8 days per month [5.0, 12.0] and a median headache intensity score of 7/10 [5.5, 8.0]. A significant proportion of individuals (57.8%) reported experiencing multiple headache types. The headaches were predominantly unilateral (72.5%) and commonly pulsating (52.3%). Associated symptoms included photophobia (96.3%), phonophobia (89.6%), and nausea or vomiting (90.8%). Nearly all participants (90.8%) reported that headaches were aggravated by physical activity. Additionally, 39.4% experienced visual aura. The median MIDAS score was 24 [13.0, 35.5], reflecting moderate to severe headache-related disability. Table 1 summarizes the participant characteristics.

**Table 1.** Participant and Headache Characteristics (N = 109)

| Participant Characteristics |  |  |
| --- | --- | --- |
| Age | 35 | [26.0, 46.0] |
| Sex |  |  |
| Male (%) | 7 | (6.5%) |
| Female | 102 | (93.5%) |
| Race (%) |  |  |
| American Indian or Alaska Native | 3 | 2.8% |
| Asian | 9 | 8.3% |
| Black | 6 | 5.5% |
| White | 91 | 83.5% |
| Hispanic | 11 | 10.2% |
| Marital Status (%) |  |  |
| Divorced | 1 | 0.9% |
| Married | 44 | 40.7% |
| Separated | 2 | 1.9% |
| Single | 59 | 54.6% |
| Widowed | 2 | 1.9% |
| Headache Characteristics |  |  |
| Headache Frequency | 8 | [5.0, 12.0] |
| Multiple Headache Types | 63 | 57.8% |
| Headache Intensity | 7 | [5.5, 8.0] |
| Location |  |  |
| Unilateral | 79 | 72.5% |
| Bilateral | 29 | 26.9% |
| Pulsating Quality | 57 | 52.3% |
| Nausea or Vomit |  |  |
| Never | 10 | 9.2% |
| >= Sometimes | 99 | 90.8% |
| Photophobia |  |  |
| Never | 4 | 3.7% |
| >= Sometimes | 105 | 96.3% |
| Phonophobia |  |  |
| Never | 11 | 10.4% |
| >= Sometimes | 95 | 89.6% |
| Aggravated by activity |  |  |
| Never | 8 | 7.5% |
| Sometimes | 99 | 90.8% |
| Visual Aura | 43 | 39.4% |
| MIDAS Total | 24 | [13.0, 35.5] |

Values are frequency counts (%) or median [25<sup>th</sup>, 75<sup>th</sup>]

Although N = 109 individuals were enrolled, only N = 104 provided diary entries. These participants self-reported 187 different headache triggers for up to 28 days, resulting in 540,876 headache trigger measurements. In total, 5,886 diary entry occasions were observed, with 2,880 AM entries and 3,006 PM entries. Of the eligible entries, 710/5,886 (12.1%) were missing, yielding a diary adherence rate of 88% for active participants.

### Information Entropy in Common Headache Triggers

Entropy provides a quantitative measure of uncertainty or variability in a system. It is calculated as the expected value of surprisal, which reflects the degree of surprise associated with observing a particular outcome. When extended across a range of outcomes (e.g., levels of a headache trigger), entropy captures the average level of unpredictability in the data. Applied to self-reported measures, entropy reveals the diversity of participants’ experiences, offering insights into the consistency or variability of specific behaviors, symptoms, or states. Higher entropy suggests greater variability or unpredictability across participants, while lower entropy indicates more uniformity or consistency in responses. This approach allows for a nuanced understanding of the experience of headache triggers, facilitating comparisons across individuals in trigger domains and highlighting within-person patterns of variability in participants’ experiences.

Figure 1A displays the entropy of sleep-related variables, reflecting the variability in participants’ self-reported experiences. Entropy values ranged from near 0 to over 4.0 bits, with notable individual differences across sleep variables. “Nap or doze” (i.e., the self-report of a nap on a particular day) exhibits the lowest entropy, indicating high predictability or uniformity in responses (i.e., individuals either always nap or never nap). Variables such as “quality of sleep,” “rested and refreshed,” and “up during the night” show moderate entropy, suggesting a mix of consistency and variability in participants’ experiences. Higher entropy is observed for variables like “time awake,” “bedtime,” and “awake time,” reflecting greater diversity and unpredictability in these measures. The figure highlights distinct patterns in the variability of sleep-related behaviors and perceptions, emphasizing differences in how participants report various aspects of their sleep.

**Figure 1.**
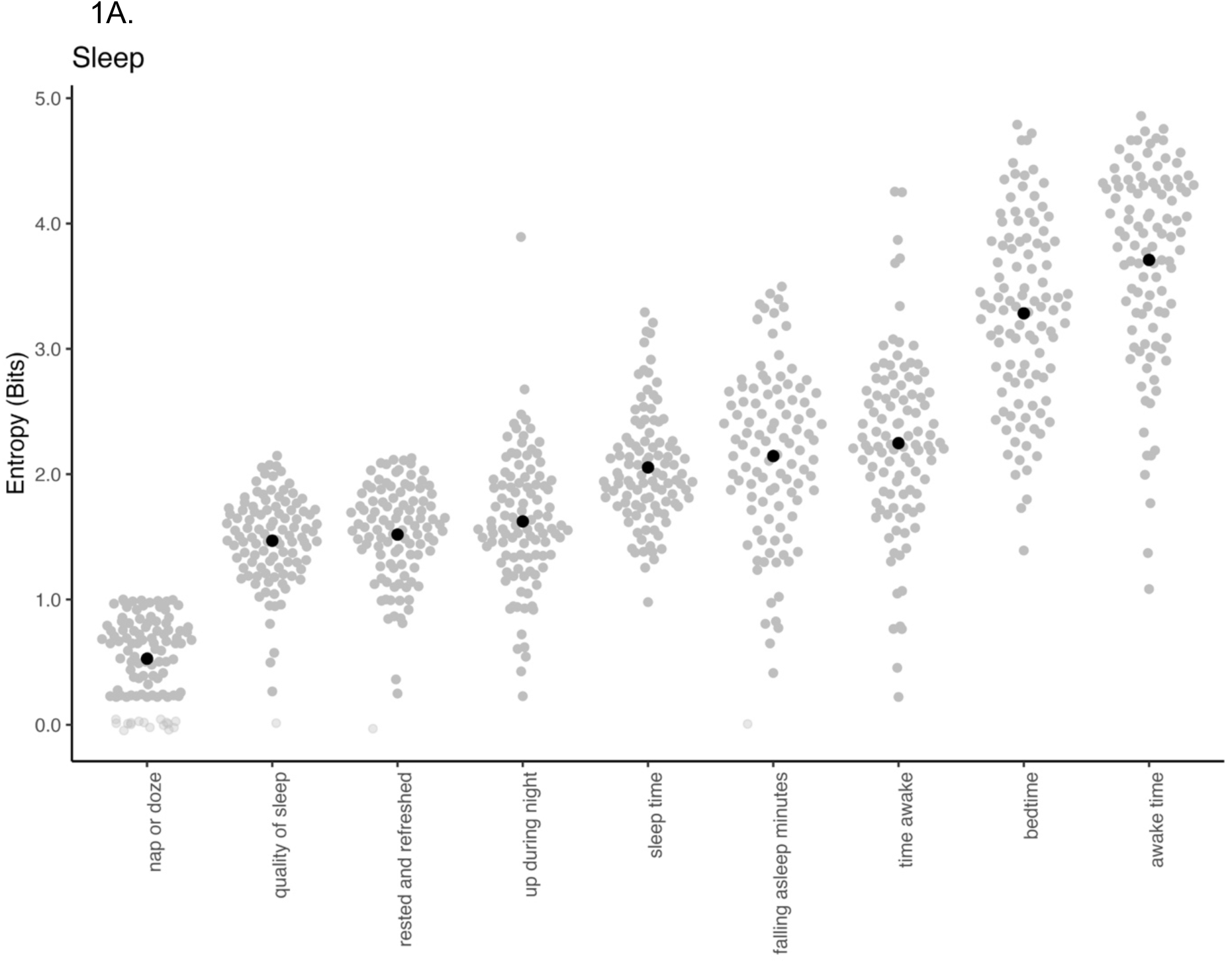
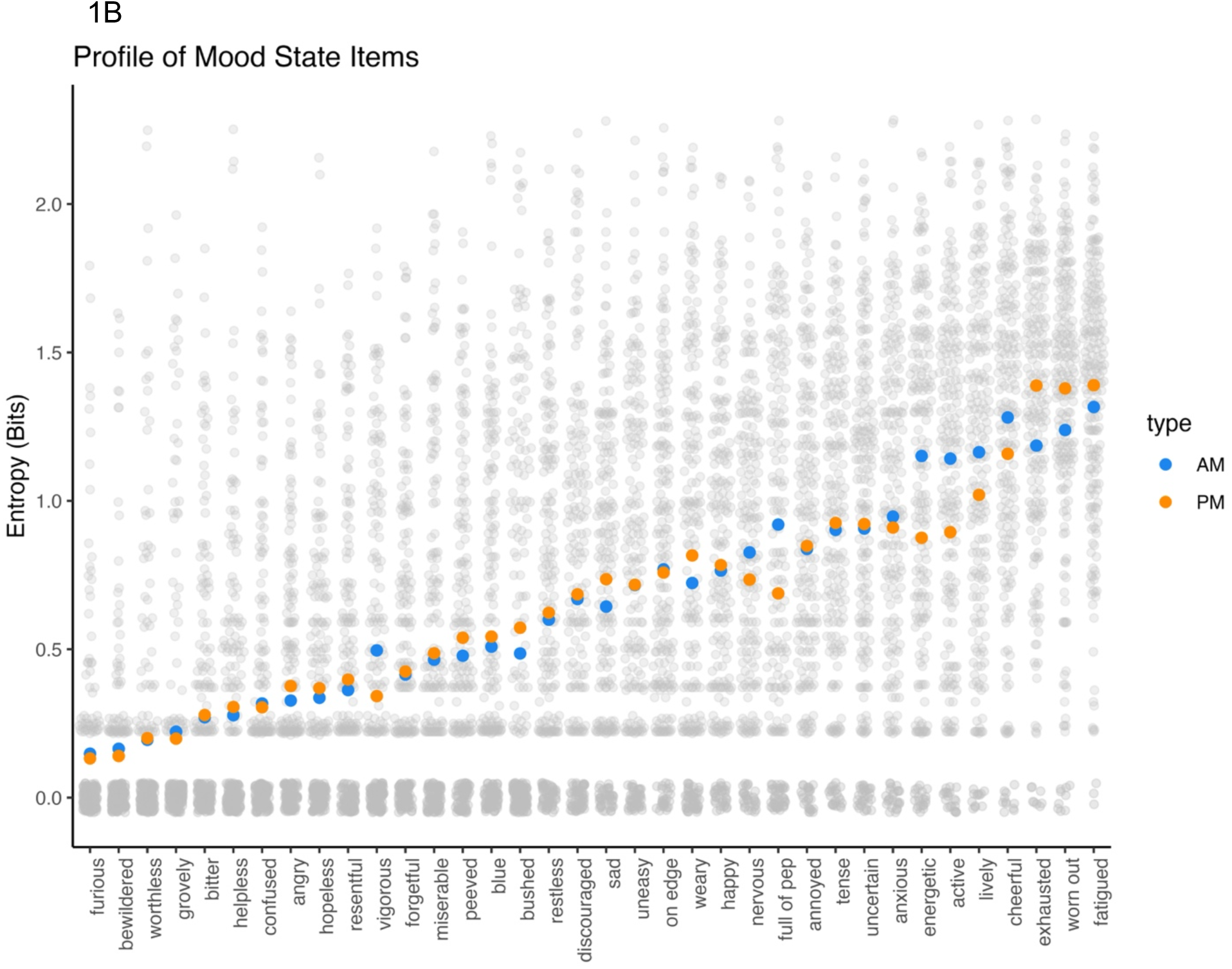
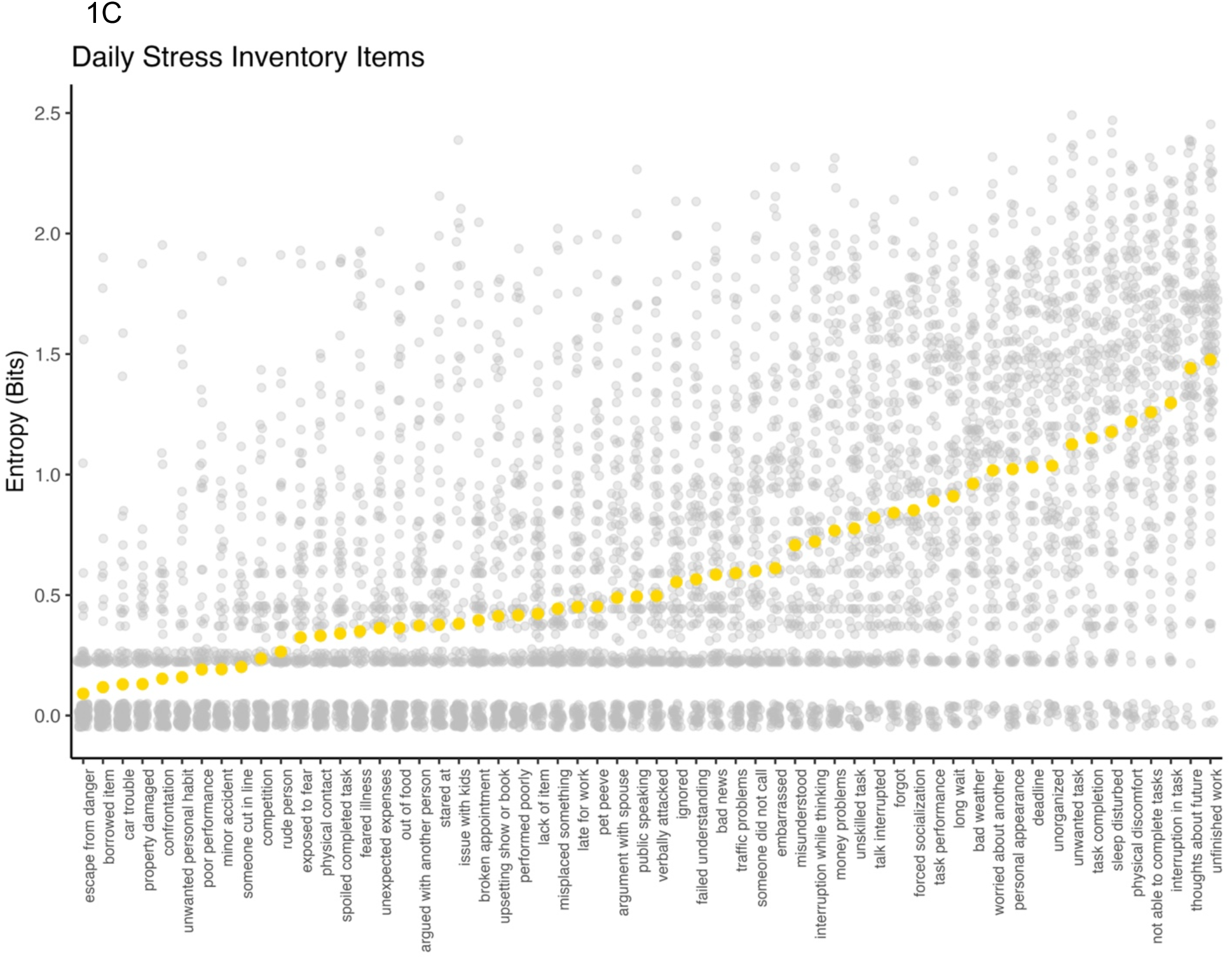
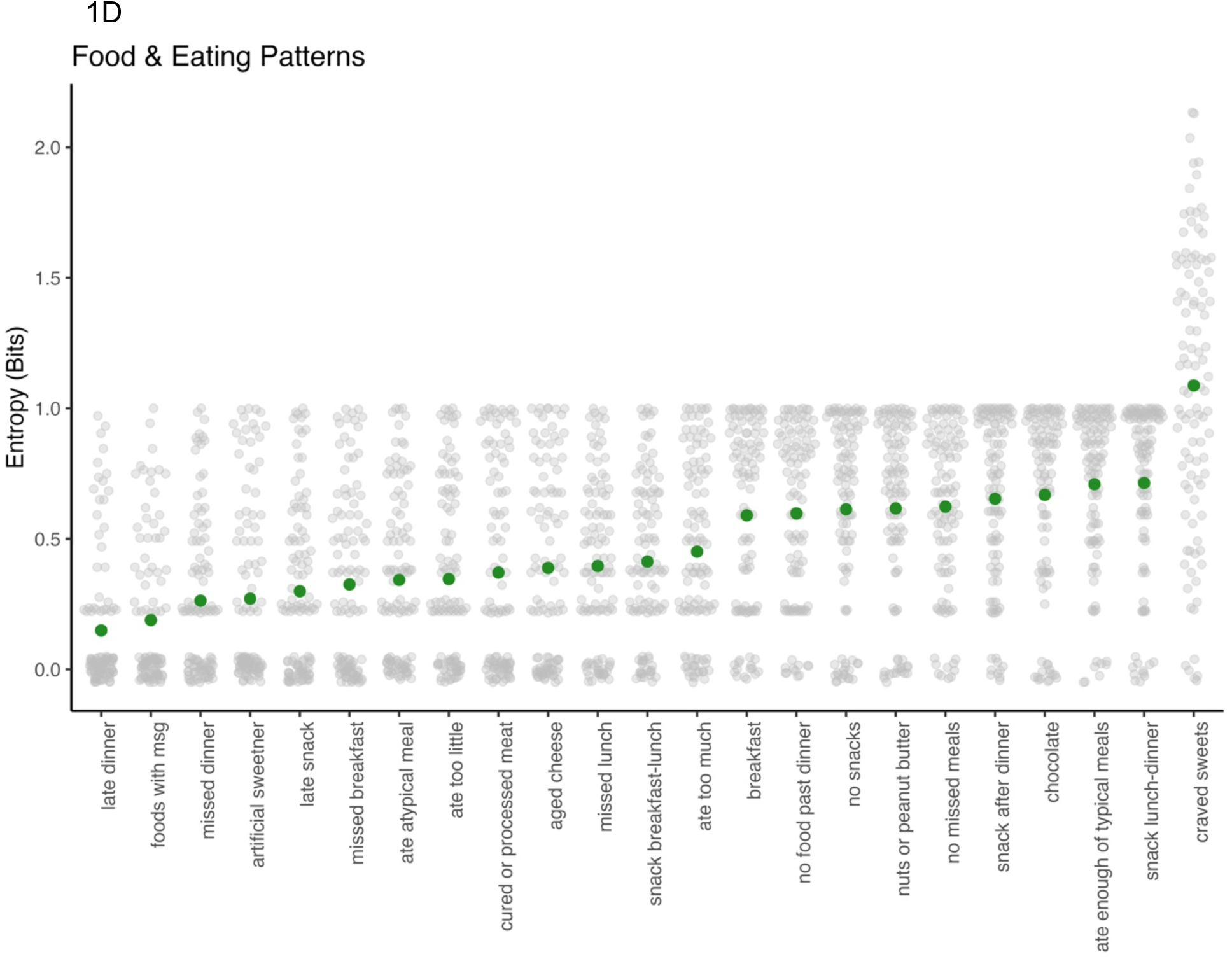
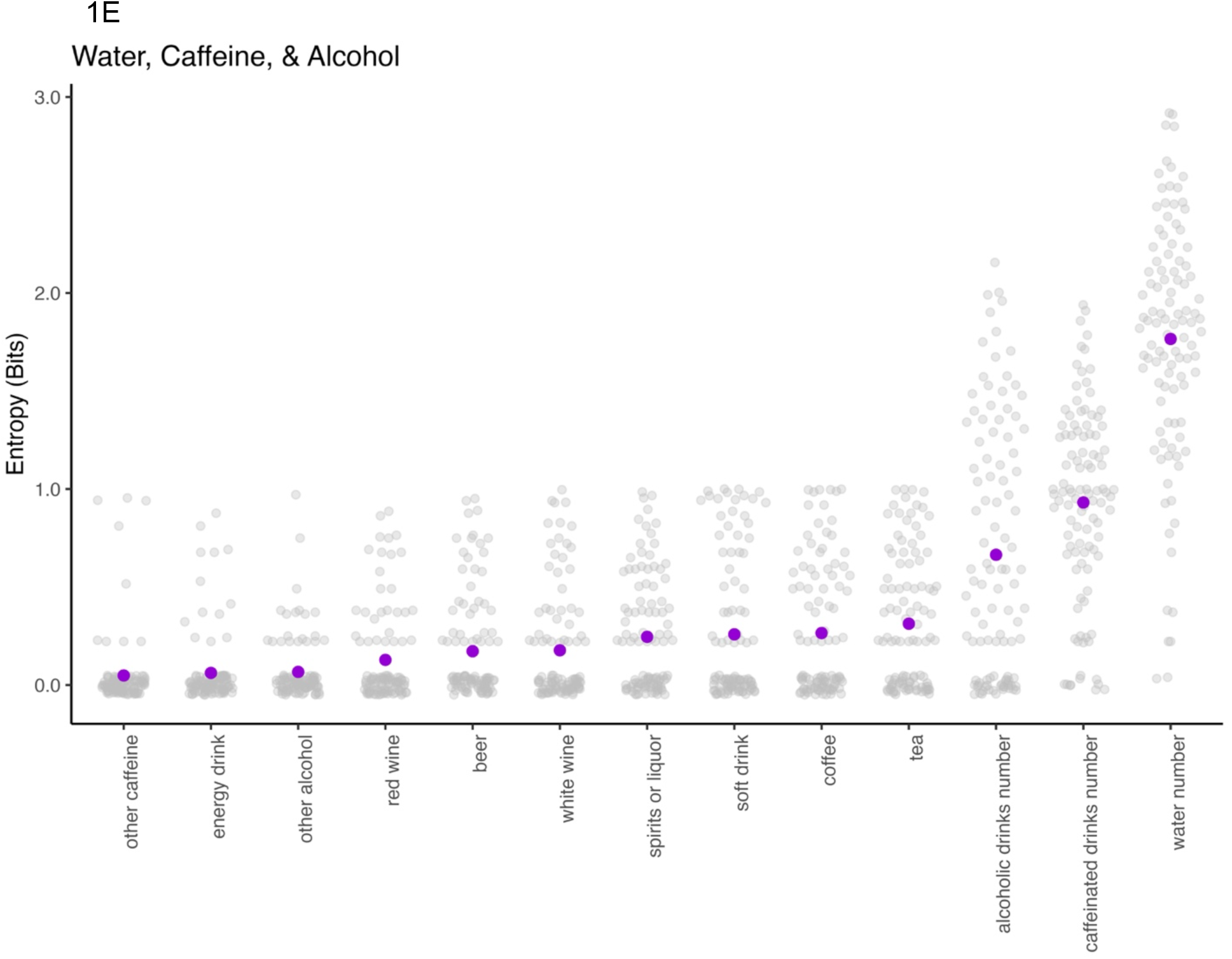
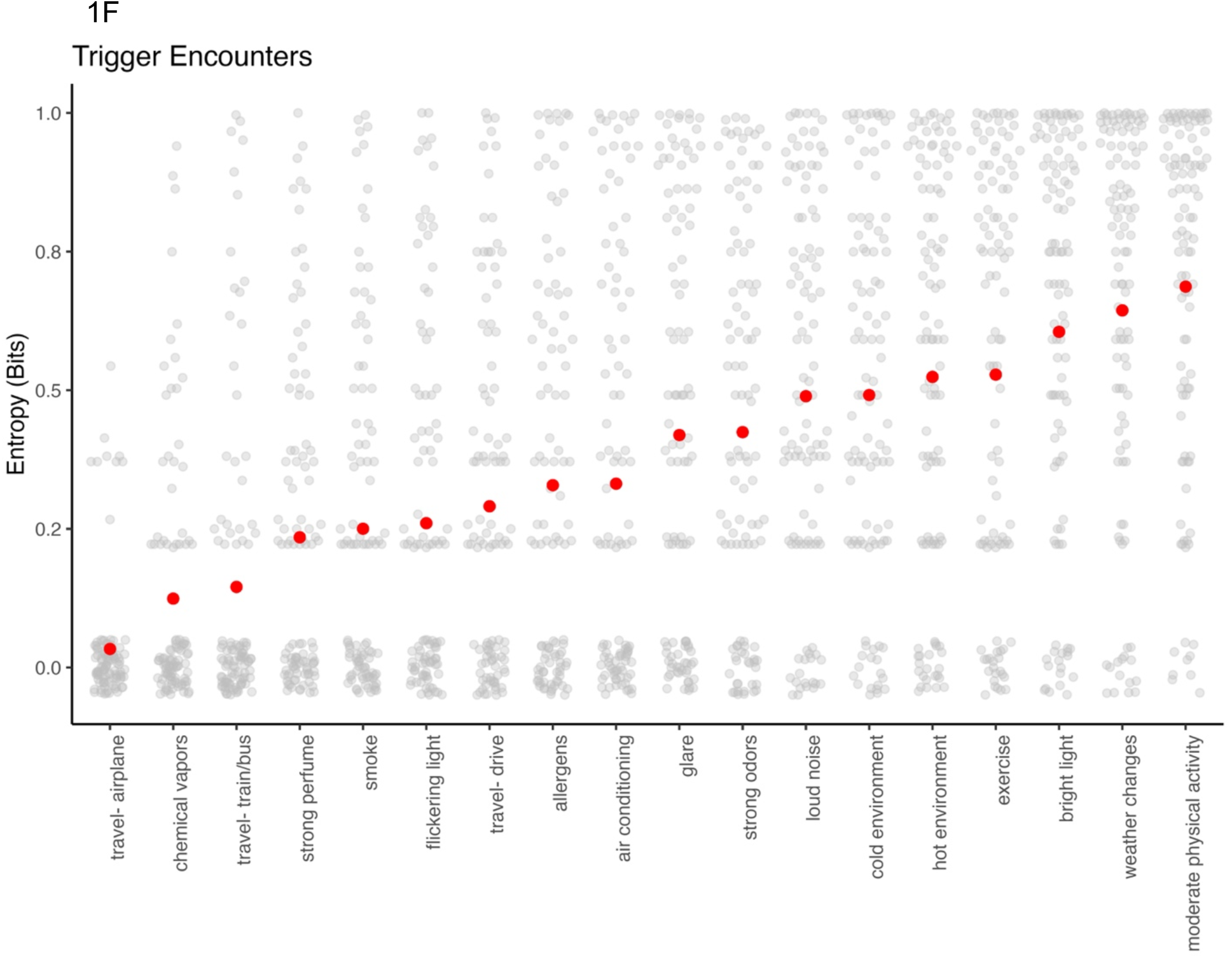
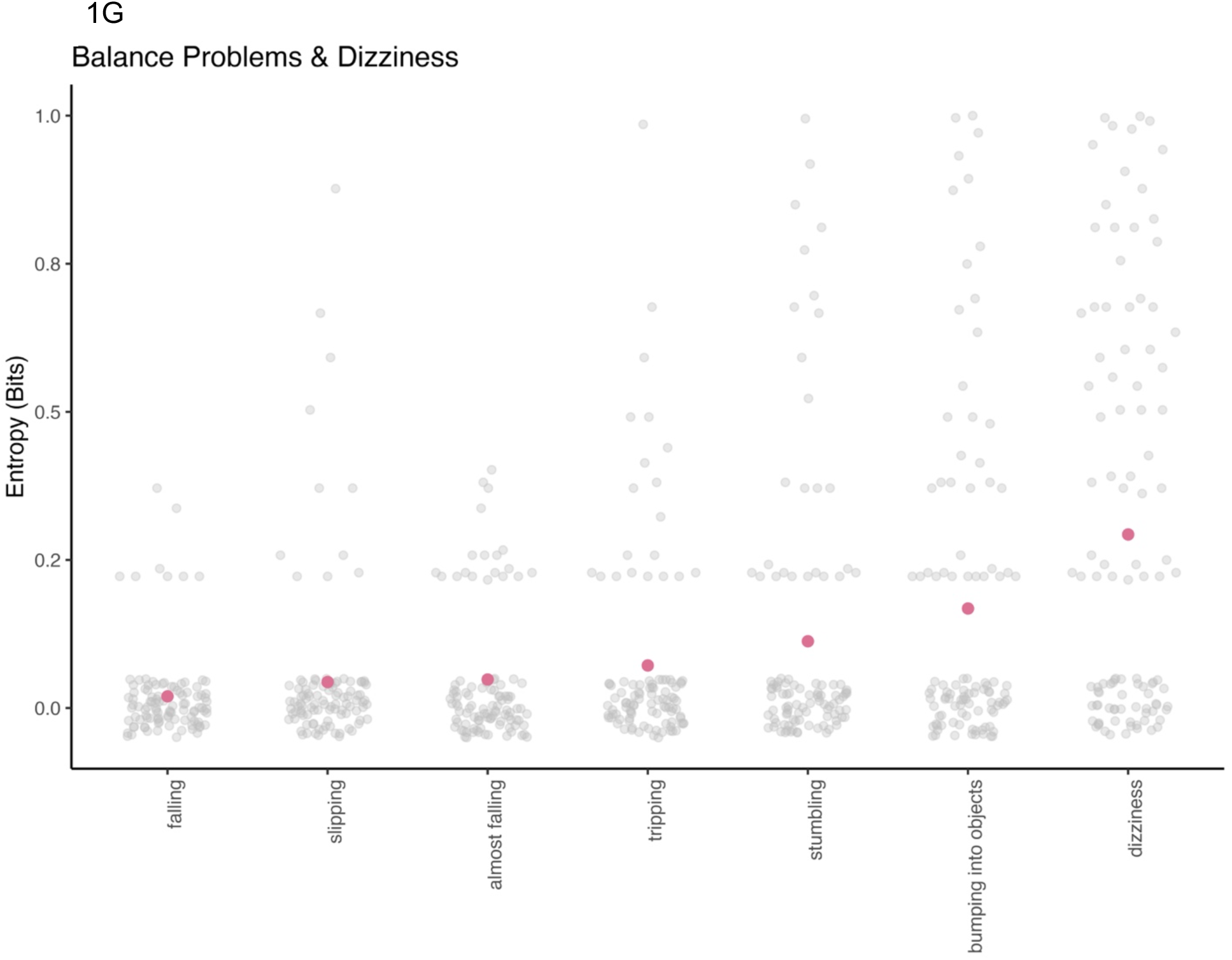
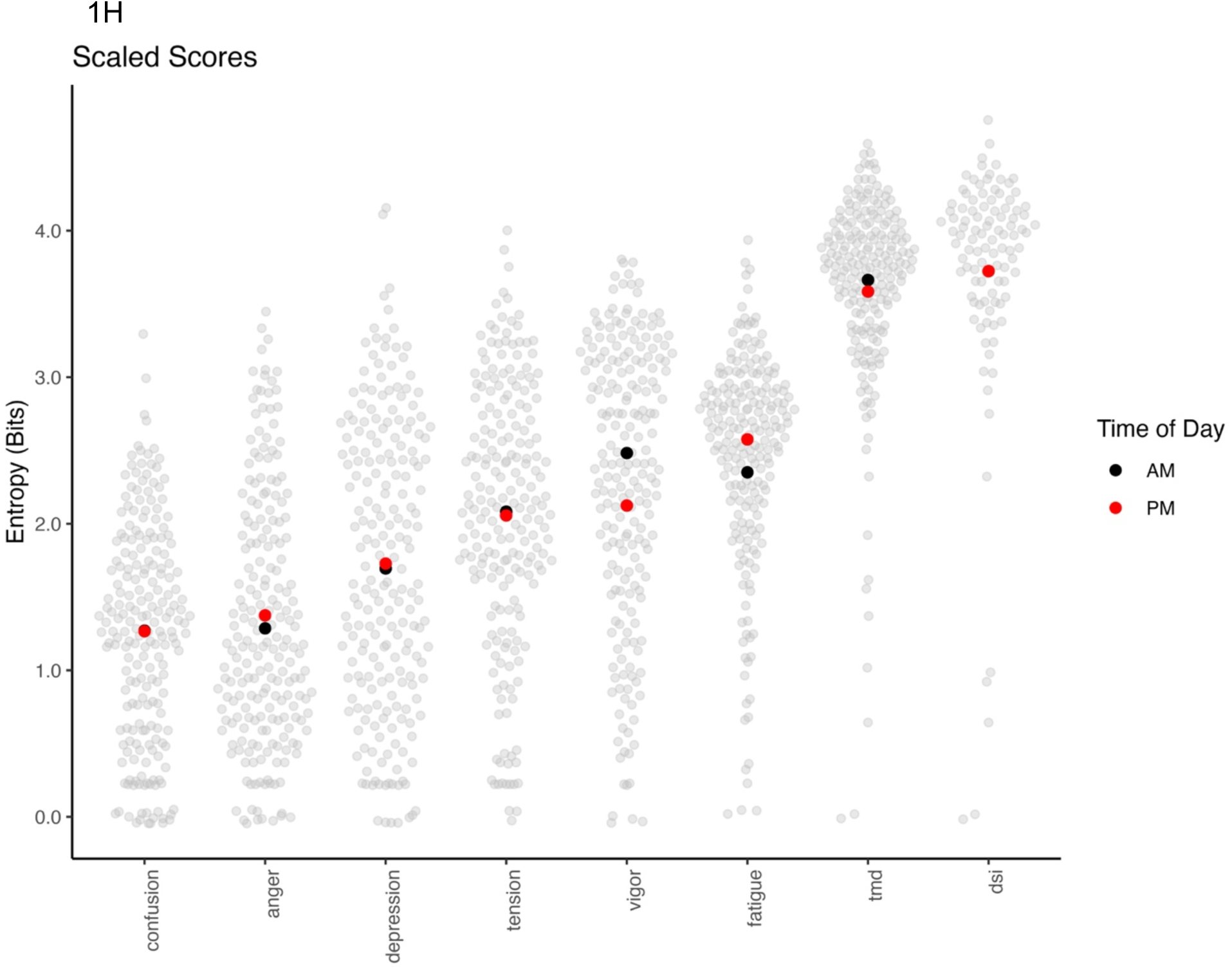
Profiles of entropy (measured in bits) across various self-reported measures. (A) The entropy of sleep-related items, including nap duration, sleep quality, and timing measures, illustrates response variability. (B) The entropy of Profile of Mood State items (POMS-SF), separated by morning (AM) and evening (PM) responses, highlighting differences in emotional variability between times of day. (C) Entropy of daily hassles from the Daily Stress Inventory (DSI). (D) The entropy of food and eating patterns such as missed meals and snacking. (E) The entropy of drinking behaviors for caffeine, water, and alcohol. (F) The entropy of common headache trigger exposures such as transportation experiences, weather, physical activity, etc. (G) The entropy associated with balance problems, dizziness, or tripping and falling. (H) The entropy associated with the mood subscales of POMS-SF, including Total Mood Disturbance (TMD), and the sum of stress items, including the total score DSI. Each point in each plot represents one individual’s data, with colored points showing item means. The between-person spread in points within item reflects the variability in reported experiences across individuals.

Figure 1B displays the entropy of mood state items (POMS-SF), categorized by time of assessment (AM vs. PM). The entropy values range from approximately 0.0 to over 2.0 bits, with lower entropy observed for items such as “furious” and “bewildered,” indicating low variability or high predictability in responses. Conversely, higher entropy is seen for items like “fatigued” and “worn out,” reflecting greater variability or less predictability. For most items, entropy values are similar across AM (blue) and PM (orange) assessments, suggesting consistent response variability regardless of the time of day. However, differences for certain items indicate potential time-of-day effects on response patterns. Positive mood states (e.g., “energetic,” “lively”) and fatigue-related states (e.g., “fatigued,” “worn out”) exhibit higher entropy, suggesting diverse daily responses, whereas negative or extreme mood states (e.g., “furious,” “helpless”) generally show lower entropy, reflecting more consistent reporting.

Figure 1C displays the entropy of items from the Daily Stress Inventory, reflecting the variability in participants’ responses to daily stressors. Entropy values range from near 0 to over 2.5 bits, with clear differences across items. Low entropy is observed for items such as “escape from danger” and “exposure to threat,” indicating these stressors are less frequently reported across participants. In contrast, items such as “unfinished work” and “thoughts about tasks” exhibit higher entropy, reflecting greater diversity and unpredictability in participants’ experiences with these stressors. The gold markers highlight the median entropy values for each item, demonstrating an overall trend of increasing entropy for these more commonly experienced stressors.

Figure 1D illustrates the entropy of food and eating pattern items, reflecting the variability in participants’ dietary behaviors. Entropy values range from near 0 to over 2.0 bits, with notable variability across different eating patterns. Items such as “late dinner,” “foods with MSG,” and “missed dinner” exhibit low entropy, indicating consistent or predictable patterns among participants for these behaviors. In contrast, items like “craved sweets” and “snack lunch-dinner” show higher entropy, reflecting greater variety and unpredictability in these dietary behaviors. The green markers represent the median entropy for each item, highlighting a general trend of higher entropy for behaviors related to snacking and cravings compared to variability in structured meal times.

Figure 1E shows the entropy of variables related to water, caffeine, and alcohol consumption, capturing the variability in participants’ drinking behaviors. Entropy values range from near 0 to over 3.0 bits, with distinct patterns across beverage categories. Low entropy is observed for items such as “other caffeine” and “energy drink,” indicating these behaviors are less variable and more predictable among participants (i.e., due to these beverages being not often encountered). In contrast, higher entropy is seen for “water number,” “caffeinated drinks number,” and “alcoholic drinks number,” reflecting significant variability in the quantity of these beverages consumed each day. The purple markers represent the median entropy for each beverage type, highlighting a pattern of increasing variability for numeric measures of consumption compared to categorical (imbibed yes vs. no) items.

Figure 1F illustrates the entropy of trigger items that involve an encounter, capturing the variability in participants’ experiences with commonly reported stimuli or experiences. Entropy values range from near 0 to approximately 1.0 bit, with most items demonstrating low entropy, indicating consistent or predictable exposure patterns. Items such as “travel-airplane,” “chemical vapors,” and “travel-train/bus” exhibit the lowest entropy, suggesting these triggers are encountered less frequently or with greater consistency among participants. In contrast, items such as “weather changes” and “moderate physical activity” show relatively higher entropy, reflecting greater diversity in how participants experience these triggers. The red markers represent the median entropy for each item, emphasizing a general trend of low variability across most trigger-related encounters, but the spread of dots indicates remarkable between-person variability in the entropy of these triggers. This figure highlights that certain environmental or situational triggers are more uniformly reported, while others exhibit greater variability in exposure both within and across persons.

Figure 1G illustrates the entropy of items related to balance problems and dizziness, reflecting variability in participants’ experiences. Entropy values are relatively low across all items, ranging from near 0 to approximately 1.0 bit, indicating consistent and predictable patterns. Items such as “falling,” “slipping,” and “tripping” exhibit the lowest entropy, suggesting these balance problems are rarely encountered among participants. Slightly higher entropy is observed for “dizziness,” reflecting somewhat greater variability in participants’ reports of this experience. This figure highlights that most balance-related issues are uniformly reported, with minimal differences in how participants experience these problems.

Figure 1H presents the entropy of subscales and total scores related to mood states (Figure 1B) and stress (Figure 1C), comparing the variability between morning (AM) and evening (PM) assessments. Entropy values range from approximately 1.0 to over 4.0 bits, reflecting notable variability across different scales. Subscales such as “confusion,” “anger,” and “depression” show moderate entropy, with similar values between AM (black) and PM (red) assessments, indicating consistent variability across times of day. Higher entropy is observed for scales like “fatigue” and “vigor,” suggesting greater diversity in participants’ responses. Due to their being a composite of the other items, the total mood disturbance (TMD) and daily stress inventory (DSI) scores exhibit the highest entropy, reflecting substantial variability in overall mood and stress levels.

### Distribution of Observed Surprisal Scores

Figure 2 illustrates the distribution of total surprisal values across the diaries in the dataset, both overall (left panel) and within n = 20 randomly selected individuals (right panel). The left panel shows a histogram of the distribution of individual total daily surprisals, revealing a highly right-skewed multimodal pattern where the majority of values are concentrated at lower surprisal levels (0–2 bits), reflecting diary entries with more predictable or common combined daily experiences. A smaller mode appears at moderate surprisal levels (2-7 bits) where a pattern of daily experiences is less likely encountered (i.e., the joint combination of these experiences is atypical for this person). Higher surprisal values (e.g., 7–10 bits), corresponding to very unusual daily experiences, occur less frequently.

**Figure 2.**
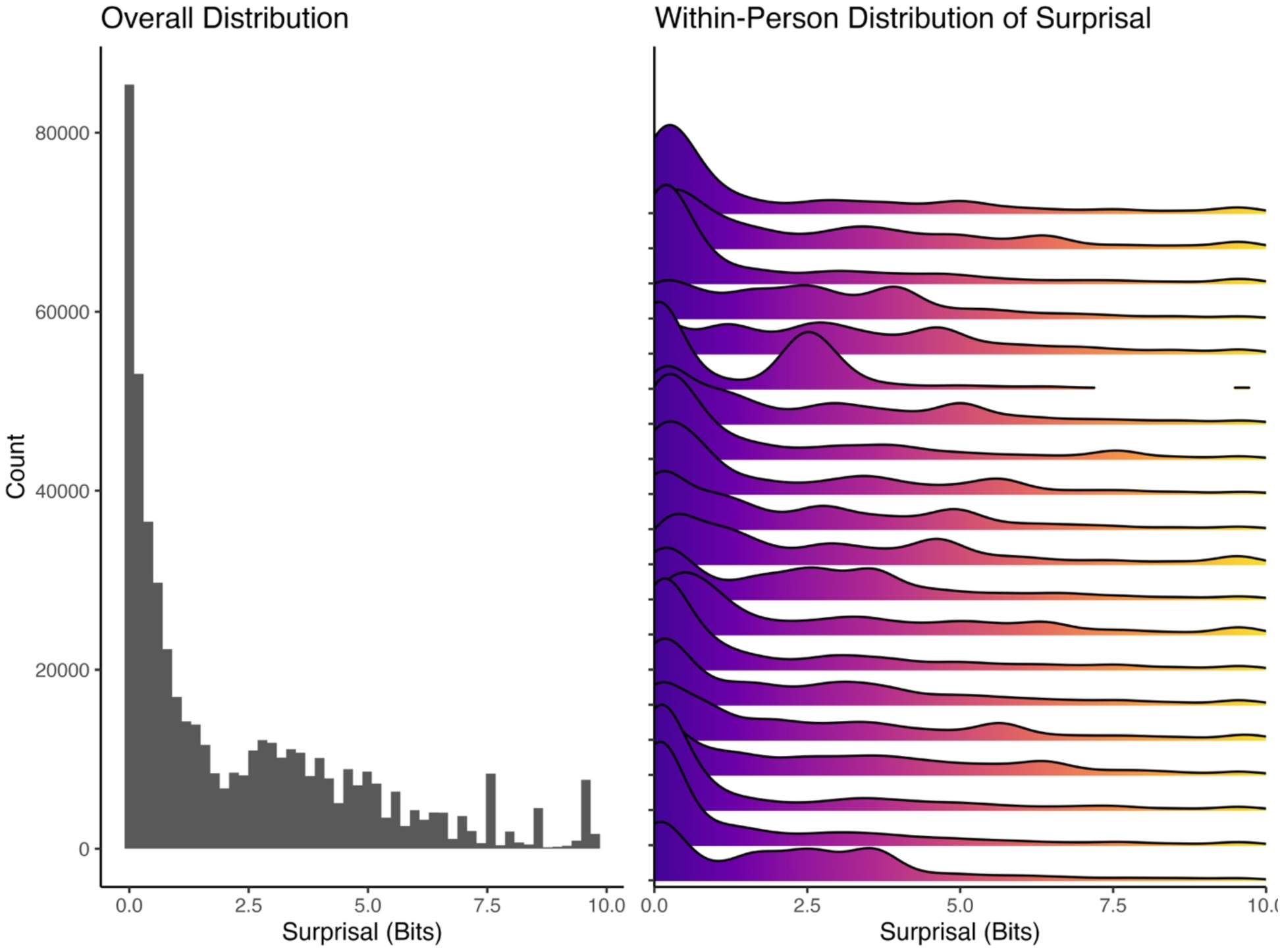
Distributions of surprisal (measured in bits) across all participants. (A) The distribution of all surprisal values across all items (N = 540,876), pooled across individuals (N = 109), shows a heavily right-skewed pattern, with most values concentrated at lower surprisal levels (0-2 bits), but with considerable days that represent a surprising combination of headache triggers. Two bits of surprise represent a combination that only occurs once every 4 days, while 10 bits are a combination that occurs only once every ∼1000 days. (B) Within-person distributions of surprisal, visualized as density ridges for n = 20 randomly selected individual participants, reveal substantial variability in surprisal patterns across individuals, with some exhibiting multimodal distributions. These results highlight the diversity in daily surprisal across and within individuals over time.

The right panel presents ridge plots of the within-person surprisal distributions, highlighting the within-person variability observed across individuals. While most participants’ distributions are similarly right-skewed, there are notable individual differences in the density and range of surprisal values. Some participants exhibit relatively narrow distributions with low surprisal values, suggesting highly consistent responses across days and that they are rarely surprised by the joint combination of headache triggers. In contrast, others show broader distributions, indicating greater variability, or days with a higher sum of daily surprisals. This figure underscores the importance for considering both overall and individual-level patterns of surprisal when creating a trigger measurement model, revealing substantial heterogeneity in the predictability of the sum of daily experiences across the sample.

### Correlation of Individual Daily Surprisals

Figure 3A presents the AM surprisal correlation matrix, highlighting the relationships between variables measured in the morning. The matrix primarily shows positive correlations (green), with stronger associations observed between related constructs, such as sleep-related variables (e.g., “quality of sleep,” “rested and refreshed,” and “sleep time”) and mood states (e.g., “fatigue,” “anger,” and “depression”). Notably, “time awake” correlates with several other time-related variables, including “bedtime” and “awake time,” reflecting interconnected patterns in behaviors. Weather-related variables (e.g., “weather affect health”) exhibit weaker and more scattered correlations with other domains, suggesting either poor reliability of these items or limited integration with broader constructs. The matrix captures a coherent structure where sleep, most mood states, and behavioral patterns are interlinked, providing a snapshot of morning dynamics in participants’ responses.

**Figure 3.**
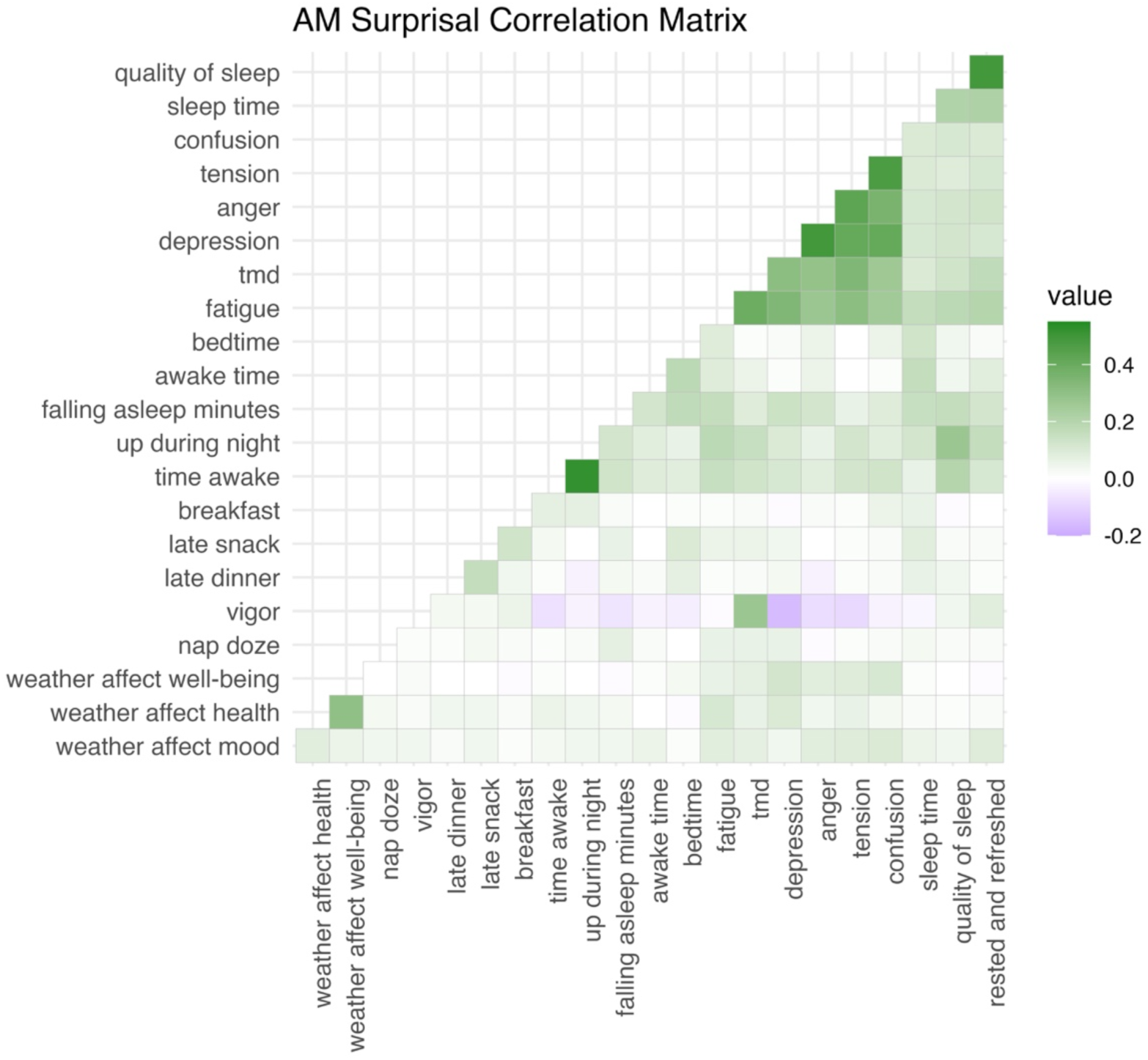
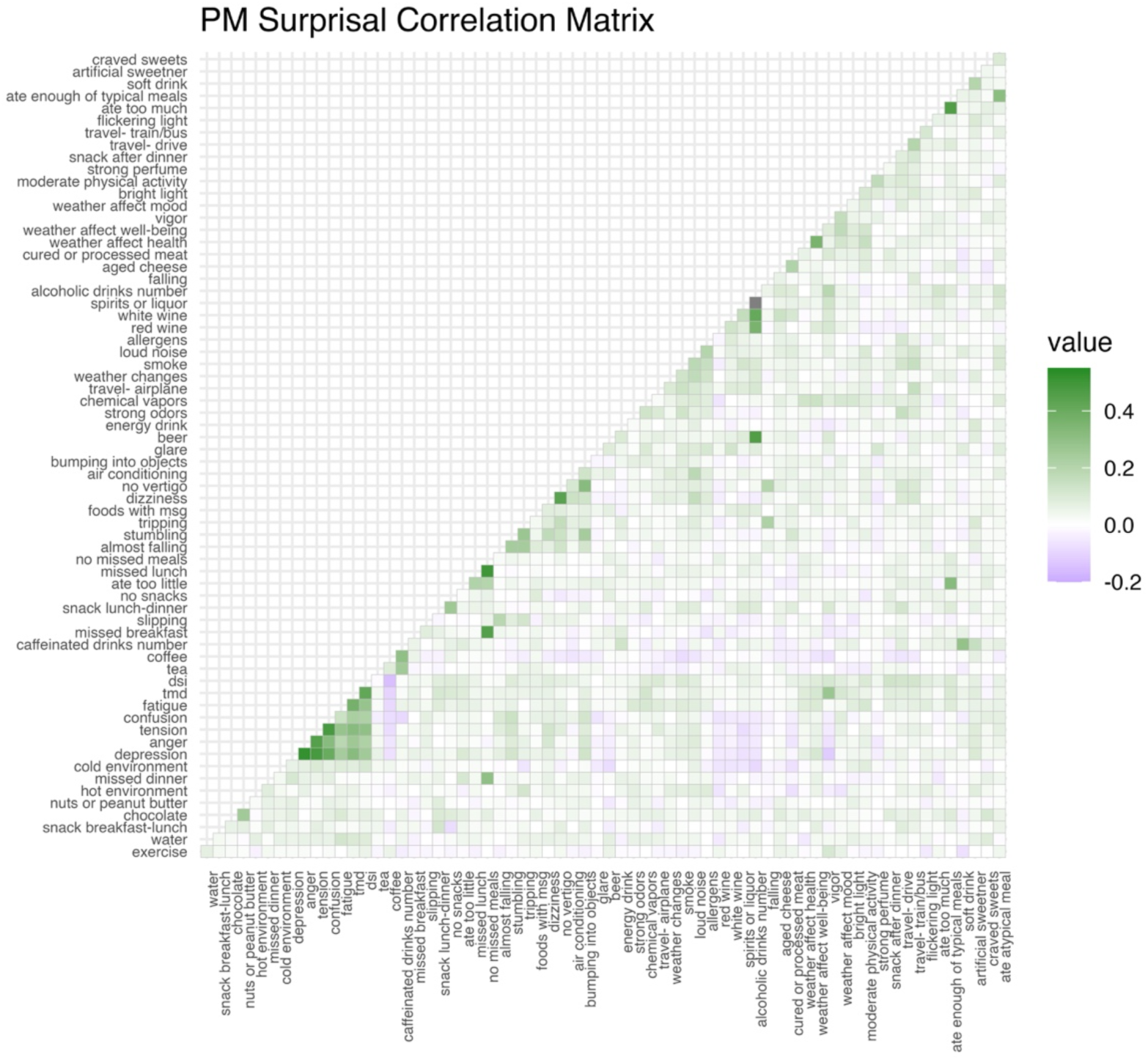
Correlation matrices for surprisal values across variables assessed in the morning (AM) and evening (PM). (A) The AM surprisal correlation matrix shows relationships between sleep-related, emotional, and behavioral variables, with stronger correlations (green-positive and purple-negative) indicated by darker colors. (B) The PM surprisal correlation matrix displays the broader range of variables assessed in the PM entry, including dietary, environmental, and physical activity factors. The patterns reflect more diverse relationships in the evening. Both panels highlight distinct clusters of variables with shared variability, providing insights into daily dynamics of surprisal across time periods.

Figure 3B depicts the PM surprisal correlation matrix, illustrating the relationships between variables measured in the evening. Compared to the AM matrix, the PM correlations appear more dispersed, with notable clusters of stronger associations. Variables such as “craved sweets,” “late dinner,” and “snack lunch-dinner” are moderately correlated, indicating patterns in evening eating behaviors. Mood variables (e.g., “fatigue,” “anger,” and “TMD”) maintain consistent relationships, similar to the AM matrix, while triggers (e.g., “strong odors,” “flickering light”) show weaker and more variable associations. The broader range of correlations in the PM matrix suggests increased variability or diversity in evening responses compared to the morning, reflecting the complexity of end-of-day behaviors and states. Together, these matrices highlight generally stable patterns across time and the evolving relationships between variables throughout the day.

### Principal Component Analysis (PCA)

Figure 4 presents scree plots from the principal component analysis (PCA) of surprisal values after removing participant-level variance created by the nesting of repeated measures within each participant. An individual PCA model was conducted separately for AM (left panel) and PM (right panel) data. For the PCA analysis, only the first part of Formula 1 was used (i.e., total surprisal while ignoring change), as changes between trigger variables are not uniform across the AM and PM entries. For example, mood, weather, and certain eating patterns are assessed in each diary (AM and PM), while stress and trigger encounters are assessed only once daily (PM only).

**Figure 4.**
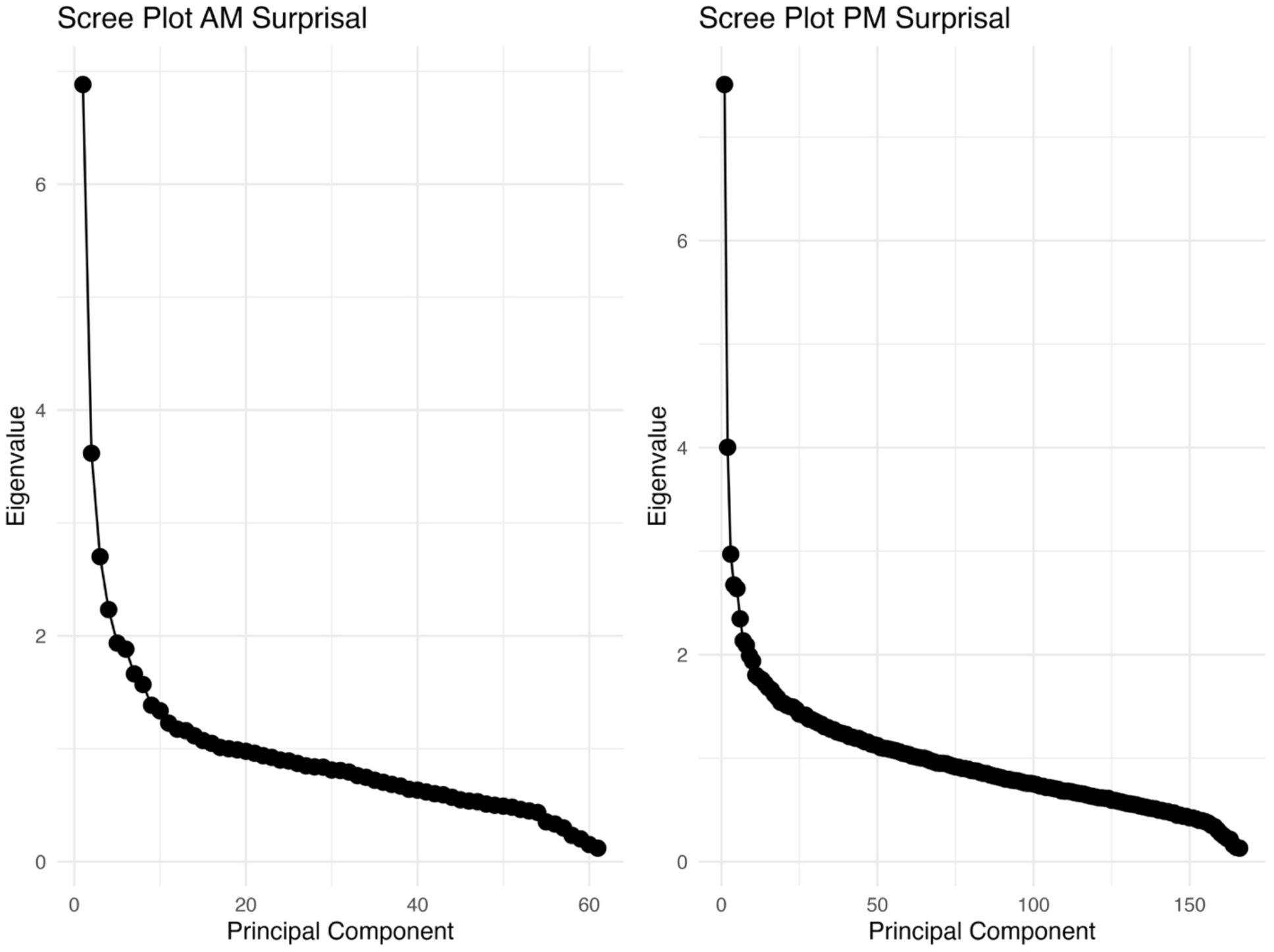
Scree plots for principal component analysis (PCA) of surprisal values in the morning (AM) and evening (PM) after removing participant-level variance from the surprisal values. (A) The scree plot for AM surprisal displays eigenvalues for the first 60 principal components, showing a steep decline followed by a leveling off, indicating that a few components explain most of the variance. (B) The scree plot for PM surprisal, which includes a larger number of variables, shows a similar pattern, with the first few components capturing substantial variance. These plots highlight the dimensionality of surprisal data across different times of the day.

Both plots display the eigenvalues associated with the principal components, illustrating how much variance each component explains. In the AM scree plot, the first few components capture a large proportion of the variance, as evidenced by the steep decline in eigenvalues for the first five components. Beyond the fifth component (28% of total variance), the curve flattens considerably, indicating that subsequent components contribute minimally to the explained variance. This suggests that a small number of components may be sufficient to summarize the primary patterns of variability in the AM data. Similarly, the PM scree plot shows a steep decline in the first few components, though the curve flattens slightly later than the AM data (20 components are required to account for 28% of the total variance). This indicates that the PM data may exhibit slightly more complex patterns, requiring more components to account for the same level of variance. In both cases, the PCA results highlight that the variability in surprisal values can be largely explained by a small subset of principal components, with slight differences in complexity between the AM and PM datasets.

## Discussion

This study explored the variability in migraine trigger experiences by applying information-theoretic measures, such as surprisal and entropy, to a diverse set of self-reported daily headache triggers. Using these measures, we aimed to quantify both within-person and between-person differences in the predictability of trigger exposures and assess their potential role in creating a universal headache trigger measurement system. Our findings highlight substantial variability in the surprisal and entropy of commonly reported migraine triggers across multiple domains, including sleep, mood, daily stressors, dietary behaviors, and environmental encounters. The results revealed that individual-level patterns of trigger exposure were highly heterogeneous, with some participants exhibiting consistent, predictable experiences and others showing broader, more variable patterns.

We also found that entropy and surprisal values varied systematically across domains and time of day. For example, AM measures of sleep and mood exhibited moderate to high entropy, reflecting a mix of consistency and unpredictability in daily behaviors. Whereas, PM assessments of mood, dietary patterns, and environmental encounters showed more complex patterns of variability, with higher entropy in some domains suggesting greater diversity in participants’ evening experiences. For measures related to behavior throughout the day, participants showed greater variability and entropy for drinking behaviors such as those quantified using counts like “water number”, “caffeinated drinks number”, and “alcoholic drinks number.” Similarly, “craved sweets” and “snack lunch-dinner”, another set of activities later in the day, showed higher entropy. The principal component analysis (PCA) further highlighted latent structures in trigger exposures, with a small number of components explaining much of the variability in both AM and PM headache trigger exposures. These findings reinforce the utility of surprisal measures for capturing nuanced patterns in the vast array of headache trigger data and support their potential as a measurement tool for stratifying trigger exposure either at the day level (e.g., today has more unexpected trigger exposures than yesterday) or individual level (e.g., individual 1 experienced more unexpected days than individual 2).

Certain trigger variables did not contain much information. The lower entropy values for measures related to balance problems and dizziness may reflect the average age of the sample population. There was a low entropy that suggested participants rarely encountered problems with “falling”, “slipping”, and “tripping” while there was a slightly higher entropy for “dizziness”.

There is no reason to expect that individuals with migraine have similar experiences or interact with similar environments. This notion was supported by the wide range of variability in each entropy plot in Figures 1A-H. If two individuals have drastically different chances of experiencing a trigger, for example, drinking tea, then it is difficult to compare the migraine risks associated with tea across two individuals using naturalistic observation.^5^ Depending on the measurement context, trigger candidates with no information could be excluded from the calculation to reduce the burden of measurement (i.e., eliminating triggers with no variability would require participants to track fewer items).

Though this analysis does not consider which triggers are associated with migraine attacks, it does provide clearer insight into how migraine trigger exposures can be measured. By considering the patterns revealed in the correlation of surprisals, a precise measurement tool that reflects trigger exposures can be created. This tool could be based on one of several assumptions. For example, summing all of the individual surprisals could create a total surprisal score that reflects the aggregate sum of unusual or unexpected events in an individual’s environment or in their responses to that environment. Such a total score would assume that each source of surprisal offers relevant information about variability in the environment and that each has equal influence. The PCA analysis provides some support for the unidimensional structure that would be required for this approach. Alternatively, a weighting scheme could be considered that differentially weights sources of surprisal, assuming that certain influences are more influential than others (e.g., stress could be weighted higher than certain exposures like perfume). A second approach to creating a measurement tool could utilize only similar sources of surprisal to create “subscale” scores. For example, surprising exposures to caffeine could be summed while ignoring any other influences. Such an approach would center on the unique insights provided by focusing only on any single source of trigger risk but might lose the comprehensiveness of the broad range of trigger variations.

This study has several limitations that should be acknowledged. First, the analysis is constrained by the finite set of headache trigger variables considered. While these variables were selected based on existing literature and clinical relevance, they do not encompass the full range of potential influences on headache occurrence. Additional measurement constructs could be incorporated in future iterations to enhance the comprehensiveness and generalizability of the system. Furthermore, the current approach does not suggest differential weights of the considered trigger variables, treating all as equally influential. However, certain triggers may have stronger or more complex effects than others, and further research is needed to determine appropriate weighting strategies. Addressing these limitations in future work will improve the robustness and applicability of predictive models for headache forecasting.

These findings reinforce the utility of surprisal measures for capturing nuanced patterns within the vast array of headache trigger data. By quantifying deviations from expected trigger exposure, surprisal provides a more refined approach to identifying potential headache-inducing factors that might be overlooked in traditional frequency-based analyses. This method enables a more individualized assessment of trigger susceptibility, allowing for the stratification of exposure at both the daily and individual levels. Such an approach holds promise for improving personalized headache management strategies, as it can help differentiate between high- and low-risk exposure periods and inform targeted interventions. Future research should explore the integration of surprisal-based measures into predictive models and clinical decision-making frameworks to further enhance their applicability in headache prevention and treatment.

## Data Availability

Data may be available upon reasonable request to the authors.

## Notes

### Competing Interest Statement

The authors have declared no competing interest.

### Funding Statement

The research reported in this publication was supported by the National Institute of Neurological Disorders and Stroke of the National Institutes of Health under award number R01NS113823.

### Author Declarations

The Institutional Review Board of Mass General Brigham gave ethical approval for this work.

